# The relationship between musicianship and pain. Is chronic pain and its management a problem for student musicians only?

**DOI:** 10.1101/2023.03.23.23287617

**Authors:** Michaela Korte, Deniz Cerci, Roman Wehry, Renee Timmers, Victoria J. Williamson

## Abstract

Neuro-biological research into chronic pain has presented reliable evidence of distinct cortical and spinal alterations in chronic pain sufferers compared to healthy individuals. Furthermore, research suggests that musicians are especially vulnerable to frequent and extended pain, supported by recent neurological investigations into musicians’ brain plasticity. However, chronic pain is not simply acute pain plus time, but a separate condition. Little is known about musicians’ emotions and chronic pain-related behaviors. Acquiring such knowledge is a crucial step in understanding how chronic pain is processed by musicians. This study investigated pain catastrophizing as one of the pain-related behaviors and emotions alongside six complementary variables: anxiety, depression, depersonalization, burnout, coping strategies and professional identity. 102 under- and postgraduate students from various higher education institutions participated in an online survey. Students were allocated to three groups according to their main study subject and type of institution: music college musicians, university musicians and university non-musicians. Group testing showed that university non-musicians’ pain catastrophizing was significantly worse compared to music college musicians.

Music college musicians and university musicians were less prone to maladaptive pain processes, despite perceiving pain for significantly longer periods of time. This novel finding indicates that chronic pain does not inevitably lead to dysfunctional pain processing for musicians and should be reflected accordingly in order to optimize pain-control. The bio-psycho-socio model of chronic pain provides the most robust framework for future research, with the aim of improving care and wellbeing provision for the management of chronic pain in musicians.

## Introduction

Music making appears on the surface to be intrinsically linked to pain and its neuropsychological impacts. Physical risk factors for the playing of various instruments, as well as for singers, have been well documented, and preventative advice was offered as early as 1700(Ramazzini 1703; Flesch 1925). Composers have invested (their) physical pain into their musical compositions as part of opera or song and/or have crafted their works to accommodate their personal injuries whilst still being able to perform at the highest level (Lorusso, Franchini, and Porro 2015; Altenmüller 2015). Thus, people have argued successfully that artists and composers who do not suffer from pain are rather the exception, whilst at the other extreme of the argument, happy individuals create nothing (Motte-Haber 2006; Freud 1918). Is this line of argument an accurate reflection of today’s musicians?

At first glance the answer might appear to be yes. Research on pain in musicians concurs that reports of pain are significantly higher in groups of musicians compared to other groups (Kenny and Ackermann 2015; Gasenzer et al. 2017). However, on closer inspection the research is focused on musculoskeletal pain perception. This is but one element in a myriad of mind and body pain research. Moreover, it has limited informative value regarding other aspects of pain, as will be shown below.

General research into pain has established two distinct concepts: acute pain, and chronic pain. Acute pain acts as a vital alert mechanism, for instance to distinguish the extent of an injury or to detect infections. It can indicate damage to the organism, but the assessment is not exclusively based only on the perception of pain, because there is no proportional relationship between the injury and the perceived pain (Kröner-Herwig et al. 2011). During a normal healing time, endogenous mechanisms such as endorphins, GABA, or monoaminergic pathways and systems all operate as preventative measures, and prevent acute pain from transforming into chronic pain.

One reason for developing chronic pain is the failure of these mechanisms. Chronic pain is understood as ‘pain that persists past the normal healing time’ of between three and six months (Merskey and Bogduk 1994). Thus, chronic pain is not the prolongation of acute pain, but a complex state of body and mind that has distinct characteristics, and is consequently understood and coded in the International Statical Classification of Diseases and Related Health Problems (ICD) as a separate condition (e.g. R52.1 or R52.2 or F45.41; depending on origin and composition of symptoms). When it is chronic, pain no longer carries any warning function, and the pain-related injury is only marginally defined by diagnosable physical damage (Bonica and Hoffman 1954). Mechanisms of chronification include neurological changes (e.g. thalamus, limbic system and cortical structures, depending on the location of the pain), emotions (e.g. depression, anxiety or burnout), and pain related beliefs and cognitions (i.e. pain-related inner monologues or meta-cognitions developing across time that lead to a modified behavior, maladaptive strategies such as learned helplessness/hopelessness or pain catastrophizing).

Based on the existing literature on musicians and pain, we can assume that the pain described here is mostly chronic. Firstly, the duration of (musculoskeletal) pain is generally noted as 12 month and longer (Gasenzer et al. 2017). Most often, this pain is due to incorrect postures. These were worse compared to non-music students and degraded significantly during music making. This is especially prominent for asymmetric instruments such as violin or flute (Ramella, Fronte, and Converti 2014). Secondly, we have neurobiological indications of pain-based changes in musicians, such as (hyper-)sensitivity of the nociceptive system and differences in cortical neuroplasticity, which are in line with pain chronification processes (Steinmetz and Jull 2013; Zamorano et al. 2023). However, there is a noticeable discrepancy between the musculoskeletal pain felt by professional orchestra musicians and the actual identifiable physical dysfunctions diagnosed by physicians in these musicians (Paap et al. 2011). While this shows evidence that a great deal of the pain perceived by musicians is chronic, based on neurobiological indicators, we have little to no evidence of how this pain is processed. This might sound minor compared to the complex neurological evidence: it is, however, an equally crucial knowledge step towards understanding the chronification process of pain in musicians.

In summary, pain prevalence is significantly higher in musicians compared to their peers and this is a process that starts as early as training (music) college. There is sufficient evidence that a great deal of the pain musicians perceive can be considered chronic. While there are few direct investigations into pain-related beliefs and cognitions, there is conflicting evidence on variables/predictors that enhance or prevent chronification (Kenny and Ackermann 2015; Voltmer et al. 2012). Moreover, due to few comparison studies of musicians’ chronic pain with other professions, there is the question of proportions: are musicians disproportionally affected by chronic pain compared to their peers and, if so, how early in their training can we assess a difference?

The present study investigated pain catastrophizing as one of the most important pain-related beliefs and cognitions in young training musicians. The results contribute to answering the following questions: (1) how does pain catastrophizing compare between student musicians and student non-musicians? (2) Do the chosen degree/course and subsequent career prospects matter when it comes to how/if pain is catastrophized? (3) What role (if any) do more general pain chronification predictors, such as depression, burnout, or professional identification with being a musician, play in the chronification process. In endeavoring to answer these questions, this study will determine if and/or how pain catastrophizing influences young student musicians and non-musicians, how or if they differ between groups, their professional identification as musicians and subsequent career prospects. To inform our choice of factors within this study, we concentrated on previously identified and well-evidenced pain chronification predictors, such as anxiety, depression, professional identity, depersonalization, burnout, coping styles and sleep. The outcomes of this study provide timely evidence on how we may better identify (music) students, who are at risk of chronic pain during their training years.

## Material and Methods

Using a predictive design and random sampling, this quantitative study observed change in variables between groups as well as predicting which combinations of variables would most likely determine a change pain catastrophizing.

### Procedure

Participants were recruited online via the students’ servers. This study was carried out in accordance with the recommendations of the university and the protocol was approved by the Ethics Committee of the Department. All participants had to be at least eighteen years of age and to study in higher education. They gave written informed consent.

#### Material

Participants were asked to share demographic details such as age, relationship status, pain perception, etc (see table 1).

**Table 1.** Demographics

|  | University Musicians | Music College Musicians | University Non-Musicians |
| --- | --- | --- | --- |
| Age (mean, SD) | 21.6; 3.44 | 27.9; 8.74 | 23.3; 7.85 |
| Family/relationship status: stable | 49% | 57% | 48% |
| Family/relationship status: single | 51% | 43% | 52% |
| Stress relief, active (running, yoga, etc.)<sup>1</sup> | 15% | 63% | 87% |
| Stress relief, other (meditation, therapies, etc)<sup>1</sup> | 67% | 9% | 7% |
| Regular practice time (years)<sup>2</sup> | 4.25 | 4.76 | 0 |
| Regular practice time (hours per day) | 2.45 | 6.14 | 0 |
| Music theory lessons (years) | 4.82 | 4.82 | 0 |
| Formal instrumental/vocal training (years) | 4.74 | 6.11 | 0 |
| Attending live concerts<sup>3</sup> | 3.22 | 5.35 | 1.05 |
| Attentively listening to music<sup>4</sup> | 30-60min | 30-60min | 0-15min |

The **Örebro Musculoskeletal Pain Screening**(Linton and Boersma 2003) measures how participants’ experience of pain affects their performance at work/higher education. In 21 questions it addresses pain beliefs and expectations. The higher the final score, the less likely the individual is to return to work and/or remain disabled by pain. The authors specified that, using a six-month prediction, 71% of patients were correctly classified (sensitivity, 72%; specificity, 70%), with a high reliability (α = .97, *p* ≤ .05) and a high internal consistency (α = .87). Total score of 105 indicate a moderate risk, ≥ 130 a high risk of being disabled by pain.

The **Hospital Anxiety and Depression Scale** (HADS) (Zigmond and Snaith 1983) collects information on depression and anxiety symptoms using two separate scales (7 items per scale) based on a 4-point Likert response (Chronbach’s α =.6). The HADS discriminates well between anxiety and depression. It is a good fit to the Rasch Model, stable across professions and less vulnerable to cultural bias. The cut-off for significant depression and anxiety was set at ≥ 9.

**T**he **Cambridge Depersonalisation Scale** (CD-9)(Sierra and Berrios 2000) covers depersonalization through 9 questions. This scale has shown adequate internal consistency and temporal stability (*α* =.92, retest reliability 10-14 days: *r*_*tt*_ = .86). Scores are added up and can reach 0 – 90, with 0 indicating no depersonalization. The cut-off point for significance was set at the level requested by the scale’s authors at ≥ 19 (short, transient) and ≥ 90 (unique condition).

The **Brief COPE** (Carver, Scheier, and Weintraub 1989) distilled the 14 scales from the original questionnaire into three scales (28 questions). This allows for diverse testing of stress coping and correlation of findings. The three scales are: active functional coping (e.g. ‘I actively did something’), functional cognitive coping (e.g. ‘I tried to find something positive in what happened.’) and dysfunctional coping strategies (e.g. ‘I used alcohol/other substances to help me through this situation’). Internal consistency was found to be good for all subscales: emotion-focused, problem-focused, and dysfunctional subscales (α = 0.72, 0.84, 0.75).

The **Copenhagen Burnout Inventory**, (CBI) (Kristensen et al. 2005) consists of three main scales: personal burnout, work burnout, and client-related burnout. The authors’ attested all three scales to have very high internal reliability (*α* = .85 - 87). This study’s design was modelled on the study Campos, Carlotto, and Marôco (2013) to reflect the dual client burnout problem of students: the client questions were doubled up, exchanging the word client with fellow student in one set, and professor in the other. These scores reflect the level of exhaustion and fatigue perceived from this interpersonal relationship that derives from the students’ interaction with fellow students and/or academic staff (professor).

The **Athletic Identity Measurement Scale** (Brewer, Raalte, and Linder 1993) is a 10-item scale that assesses the strength and exclusivity of professional identity. The higher the score, the more a candidate identifies with being an athlete (10 – 70, mean of 40; internal consistency of *r* = .93; test-retest reliability of *r* = .89. We used Vitale’s (Vitale 2009) adaptation for musicians, changing the word athlete to musicians and called the questionnaire Musicians Identification Measurement Scale (MIMS).

The **Goldsmiths Musical Sophistication Index (Gold-MSI)** (Müllensiefen et al. 2014) is a self-reported test that assesses an individual’s propensity to engage with music. It is modelled on multidimensional construct of musical sophistication. With Chronbach’s *α* = .914 the scale is suitable instrument. We used two of the test’s subscales: active engagement and musical training (7 and 9 questions).

The **Pittsburgh Sleeping Quality Index** (PSQI) (Buysse et al. 1989) measures subjective quality of sleep (9 questions). The sleep questionnaire was administered to the music college musicians only. All participants had a regular performance schedule compared to both other groups which gave rise to the idea that this variable could influence pain perception and management. It has a high test-retest reliability and a good validity for different age groups (Cronbach’s Alpha α = .69). The PSQI was able to correctly identify 88.5% of all patients and controls, representing a 89.6% sensitivity and a 86.5% specificity rate, using an empiric cut-off point of 5.

### Statistical analysis

Evaluation of the data was performed using RStudio (2022.12.0) (Team 2013) and G*Power (Faul et al. 2009). Power of β = .8 was considered as appropriate. Power calculations found the minimum for pairwise comparison with an expectation of non-linear distribution to be 96 participants in total (33 participants per group). Missing values were imputed using the package Hmisc. Rpart and rpart.plot were used for the tree model to analyse predictability. Chi square, Mann-Whitney U-test and ANOVA were used for (pairwise) comparisons. Bonferroni corrections were applied to safeguard against multiple testing, and Spearman’s correlation for correlations.

## Results

102 under- and postgraduate students (75% United Kingdom, 16% other European Union countries, 9% United States of America; age mean = 23.6 years) from various institutions and with different primary study subjects (62% music, 38% medicine, psychology, biology) participated in this study. 67 students were from the University of Sheffield and 36 students from various music colleges. The students from music colleges remained together in one group, while the group of university students was made up of two sub-groups: 31 university musicians and 36 university non-musicians. The university musicians’ group comprised students who self-identified as musicians irrespective of their main study subject (music or science). Participants in this group showed equal levels of practice, lessons taken and engagement with music as the group of students from music college (see table 1). Participants in the university non-musician group showed hardly any engagement with music (no instrumental or theory lessons). For ease of reference, the three different groups will from now on be referred to as university musicians, university non-musicians, and music college musicians (see all tables below).

### Scale Outcomes

#### Pain perception and stress relief

39% of music college musicians had perceived pain for 12 months or longer, compared to 32% of university musicians and 22% of university non-musicians. 77.7% of music college musicians and 14.7% of university musicians reported to perceive pain while playing. 2.9% of music college musicians and 40% of university musicians reported very strong perceived pain while playing that stressed them considerably. A post-hoc found that the length of pain perceived by music college musicians was significantly longer to that perceived by university musicians *t*(58) = 5.87, *p* < .001, *d* = .3 and university non-musicians *(t* (61) = 6.0, *p* < .001, *d* = .4). 77.1% of music college musicians, 82% of university musicians and 94% of non-musicians followed an active or passive stress relief management plan (e.g. yoga, meditation).

#### Pain catastrophizing

Non-musicians had a significantly higher prevalence of high pain catastrophising (≥130) compared to music college musicians and university non-musicians (z = 1.94, p= .05). Although university musicians showed a lower prevalence of high catastrophizing compared to university non-musicians (6.94%), this was not statistically significant (p = .3) (see Fig1).

**Figure 1:**
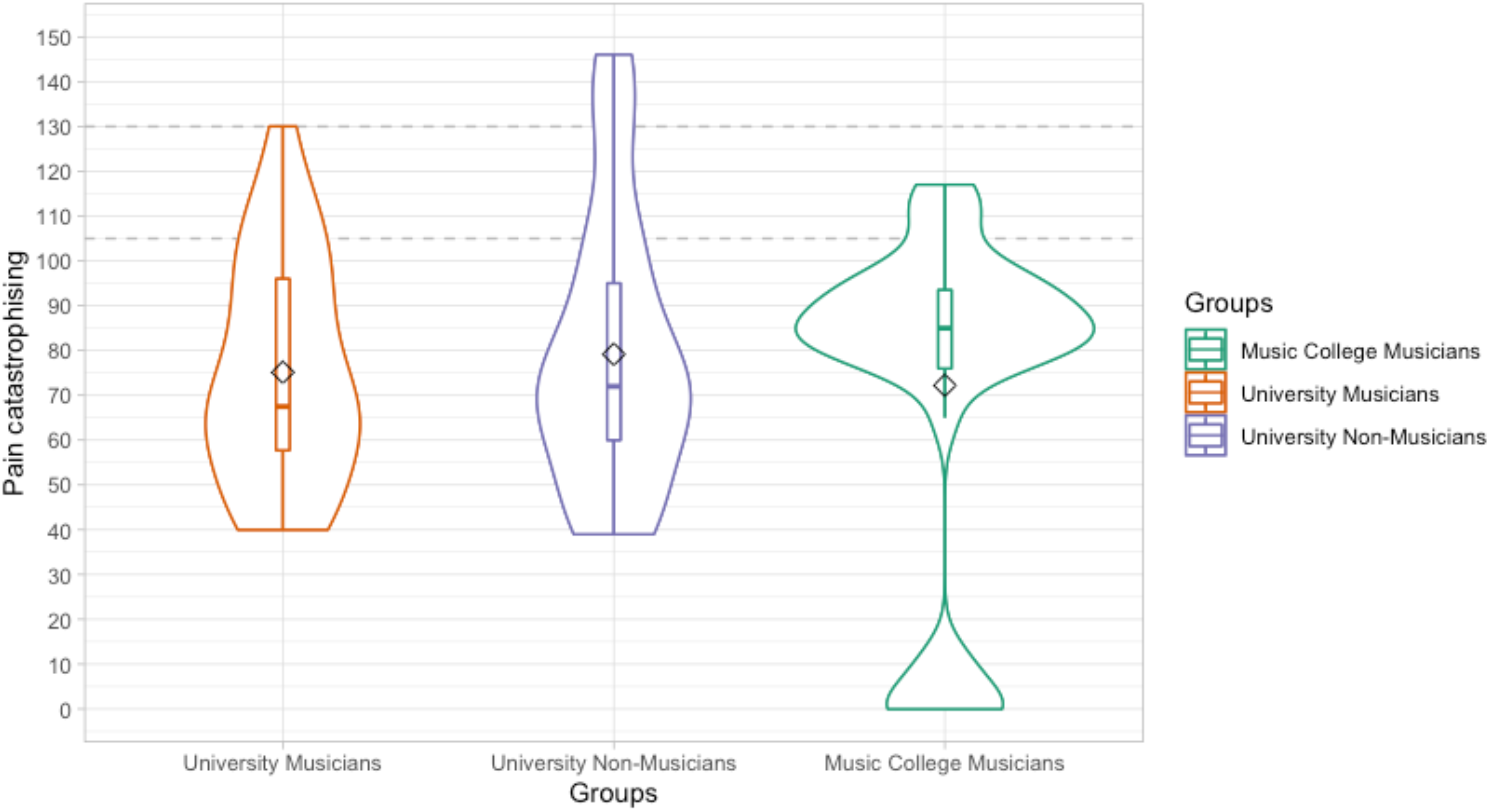
Violin plot for the Orebrö Musculoskeletal Pain Screening representing the group distribution for music college musicians, university musicians and university non-musicians, including box plot with mean points [diamond shape] and a dotted cut-off lines at ≥ 105 for moderate and at ≥ 130 for high pain catastrophising

### Depression and anxiety

There was a significant difference in depression prevalence between the music college musicians’ group and both the university musicians (*z* = -3.67, *p* = .0002), and the university non-musicians (*z* = 2.16, *p* = .003). Music college musicians showed significantly higher depression prevalence compared to university musicians and non-musicians (see table 2). The highest anxiety prevalence was found in university non-musicians. When compared to music college musicians, the difference was significant (*z* = -2.01, *p*= .04). There was no statistically significant difference between university non-musicians and university musicians (*p* = .2).

**TABLE 2.**
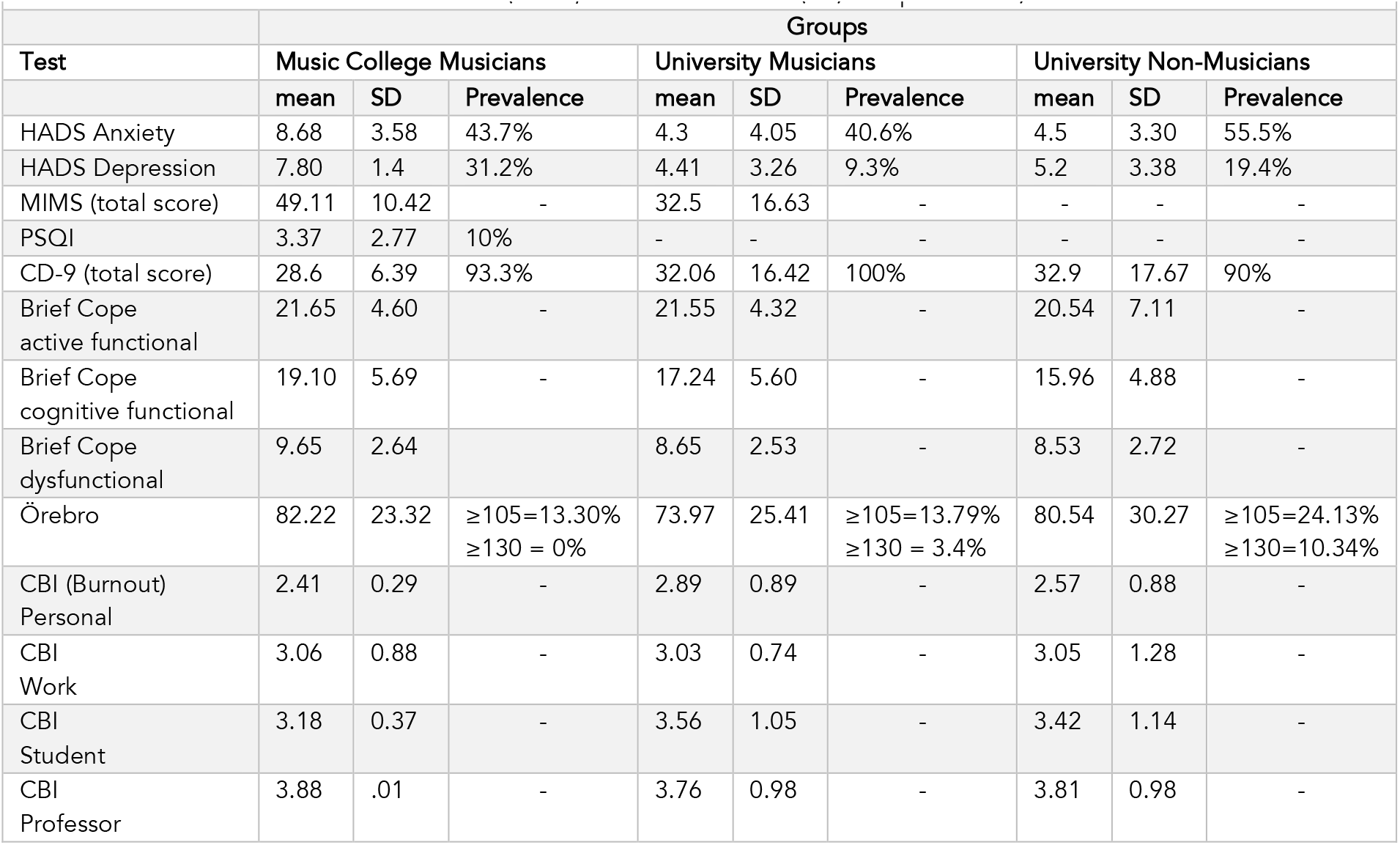
Results from all standardized tests (mean, standard deviation (SD) and prevalence)

|  |  |  |  |  |  |  |  |  |  |
| --- | --- | --- | --- | --- | --- | --- | --- | --- | --- |
| Test | Groups |  |  |  |  |  |  |  |  |
|  | Music College Musicians |  |  | University Musicians |  |  | University Non-Musicians |  |  |
|  | mean | SD | Prevalence | mean | SD | Prevalence | mean | SD | Prevalence |
| HADS Anxiety | 8.68 | 3.58 | 43.7% | 4.3 | 4.05 | 40.6% | 4.5 | 3.30 | 55.5% |
| HADS Depression | 7.80 | 1.4 | 31.2% | 4.41 | 3.26 | 9.3% | 5.2 | 3.38 | 19.4% |
| MIMS (total score) | 49.11 | 10.42 | - | 32.5 | 16.63 | - | - | - | - |
| PSQI | 3.37 | 2.77 | 10% | - | - | - | - | - | - |
| CD-9 (total score) | 28.6 | 6.39 | 93.3% | 32.06 | 16.42 | 100% | 32.9 | 17.67 | 90% |
| Brief Cope active functional | 21.65 | 4.60 | - | 21.55 | 4.32 | - | 20.54 | 7.11 | - |
| Brief Cope cognitive functional | 19.10 | 5.69 | - | 17.24 | 5.60 | - | 15.96 | 4.88 | - |
| Brief Cope dysfunctional | 9.65 | 2.64 | - | 8.65 | 2.53 | - | 8.53 | 2.72 | - |
| Örebro | 82.22 | 23.32 | $\geq 105 = 13.30\%$<br>$\geq 130 = 0\%$ | 73.97 | 25.41 | $\geq 105 = 13.79\%$<br>$\geq 130 = 3.4\%$ | 80.54 | 30.27 | $\geq 105 = 24.13\%$<br>$\geq 130 = 10.34\%$ |
| CBI (Burnout) Personal | 2.41 | 0.29 | - | 2.89 | 0.89 | - | 2.57 | 0.88 | - |
| CBI Work | 3.06 | 0.88 | - | 3.03 | 0.74 | - | 3.05 | 1.28 | - |
| CBI Student | 3.18 | 0.37 | - | 3.56 | 1.05 | - | 3.42 | 1.14 | - |
| CBI Professor | 3.88 | .01 | - | 3.76 | 0.98 | - | 3.81 | 0.98 | - |

### Depersonalization

There was almost no difference in depersonalization prevalence between college musicians (93.3%), university musicians (100%) and university non-musicians (90%).

### Coping

An ANOVA found no significant difference between groups for this variable (active functional cope: *p* = .6; cognitive functional cope: *p* = .2; dysfunctional cope: *p* = .7).

### Burnout

There was no statistically significant difference between groups. The highest level of burnout was experienced based on interactions with teaching staff, followed by fellow students, personal burnout and then work burnout.

### Sleep

With a poor sleeper prevalence of only 10%, the hypothesis that lack of sleep would have a negative impact on for music college musicians was not supported. However, there was a strong correlation between daytime dysfunction (the need to sleep during the day) and depression (r_*s*_ (29) =.91, *p* = .001). All participants with a bad sleeper score also reported high prevalence in depression, anxiety (< 9) and pain catastrophizing (< 110).

### GOLD-MSI and MIMS

Music college musicians invested more time into daily practice and formal lessons than university musicians but had not accumulated more years of practice. Music college musicians spent more time listening attentively to music and attended more live concerts (audience) than university musicians. Non-musicians took no instrumental or theory lessons. They listened less to music and attended fewer concerts (see table 2). Moving on to the MIMS, a Mann-Whitney U-test determined a significant difference in the full score between music college musicians and university musicians, with a large effect size (U = 856.0, p = .001, rank-biserial correlation = .57). The subscales self-identity (U = 853.0, p < .004) and social identity (U = 588.0, p < .004) were significantly higher. Negative affectivity (p = .4) and exclusivity (p = .5) did not differ significantly (see Fig 2)

**Figure 2:**
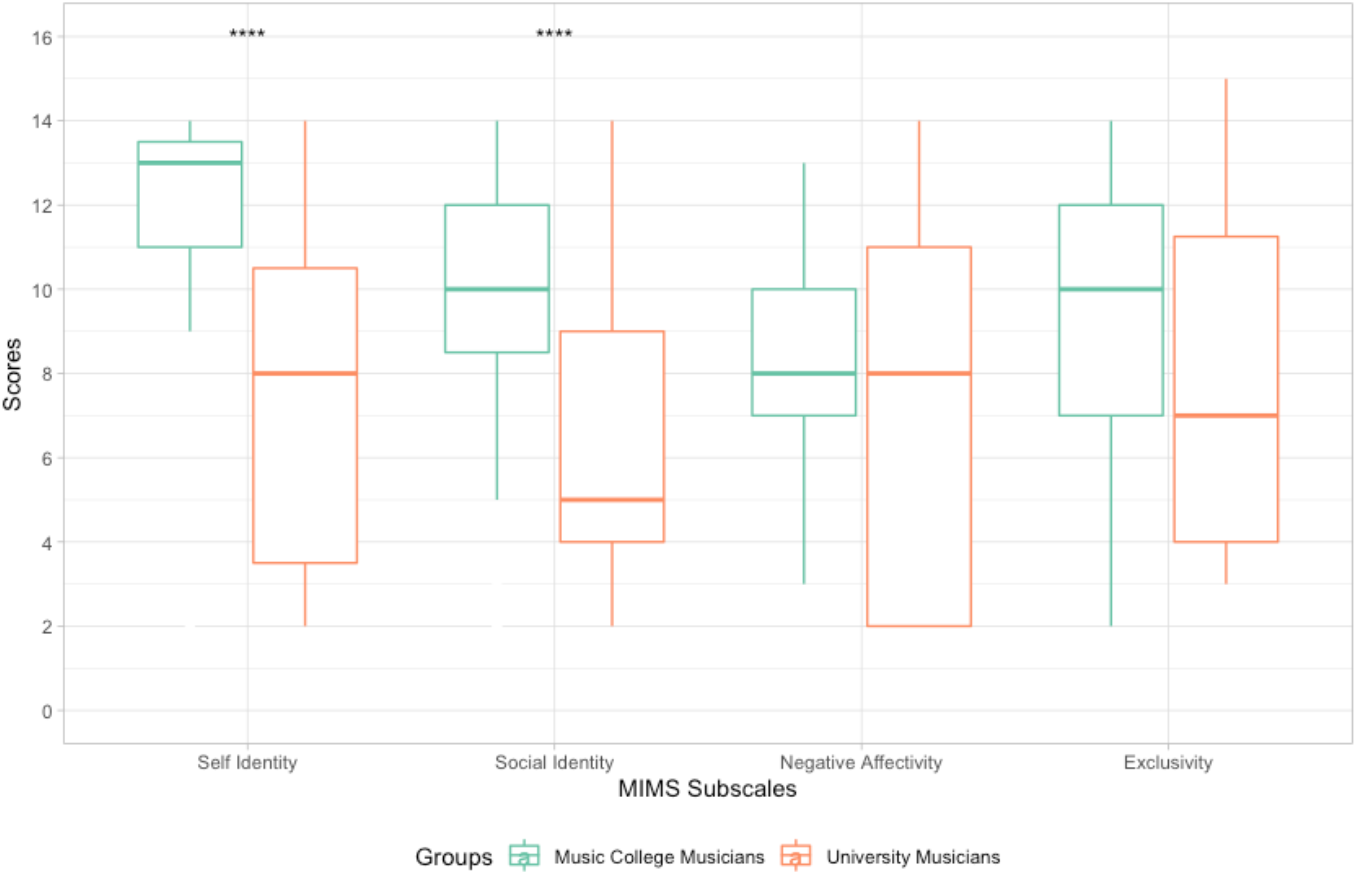
Boxplot for MIMS subscales [self-identity, social identity, negative affectivity, and exclusivity] comparing professional identity for music college musicians and university musicians; significant differences marked by asterisks

**Tree model** predicting the likeliest combination of variables that predicted a pain catastrophizing based on all variables (minus sleep) (see Fig.3)

**Figure 3.**
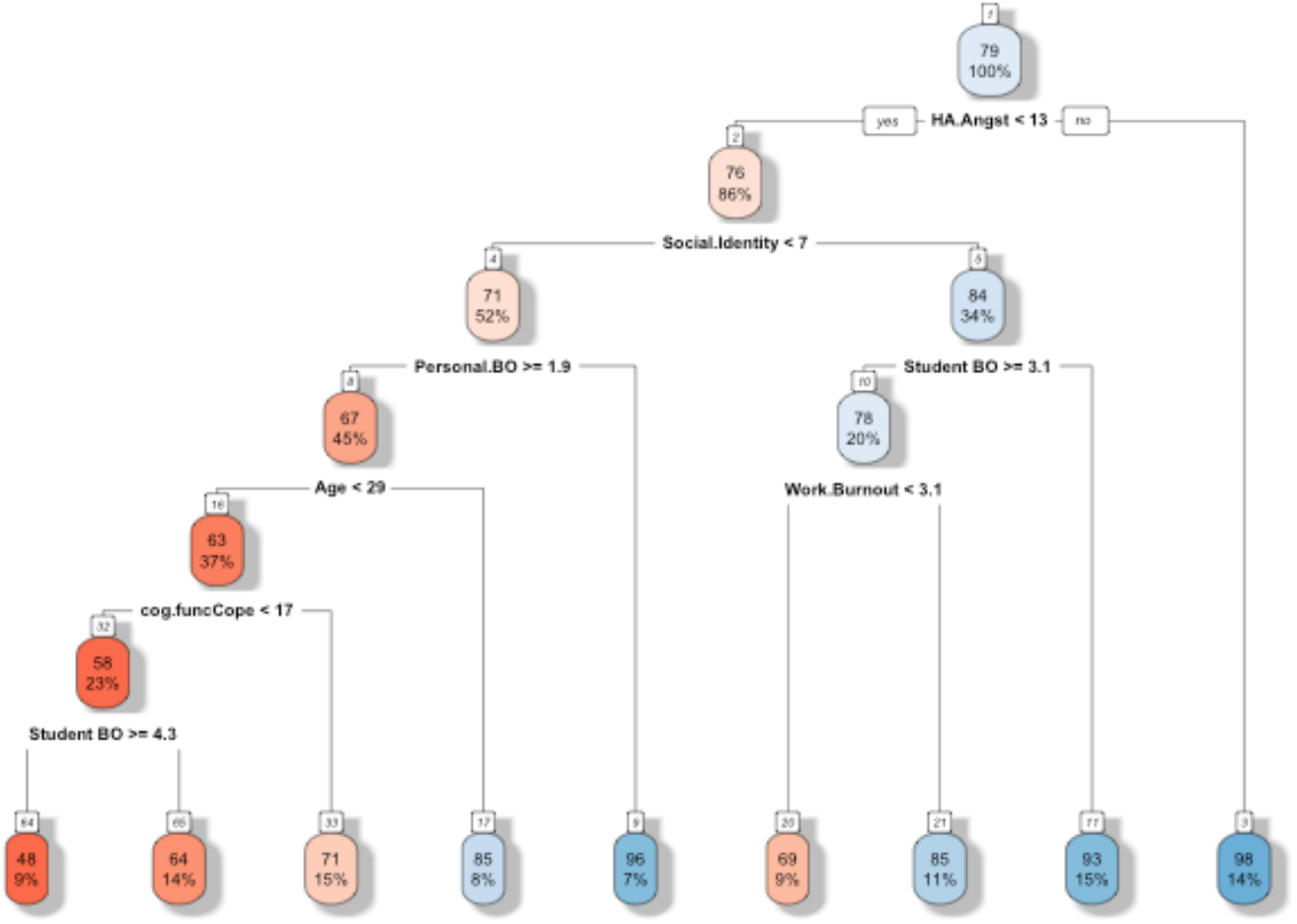
Tree model predicting the combination of variables needed for a high likelihood of pain catastrophizing. Following the red boxes from top to bottom, the highest likelihood combination (i.e. the most significant predictor pathway) is anxiety (HA_Angst: <7), social identity (<7), low personal burnout (Personal.BO: >1.9), over 29 years of age, low cognitive functional abilities (cog.funcCope: <17) and high burnout with fellow students.

The tree model illustrates that pain catastrophizing requires a combination of different cognitions/beliefs. Starting at the top box (anxiety yes/no), our models demonstrates that anxiety on its own does not lead to pain catastrophizing. The pathway to effect requires a further combination of characterized social identity (< 7), low personal burnout (> 1.9), being over 29 years of age, having low cognitive functional abilities (< 17) and high experience of burnout in relation to fellow students.

In summary, non-musicians in our study were significantly more affected by high pain catastrophizing compared to music college musicians and university musicians, despite music college musicians perceiving pain for the longest (12 month and longer) compared to both other groups. Two thirds of music college musicians reported pain while playing, compared to less than one sixth of university musicians. However, only 2.9 % of music college musicians reported very strong pain while playing that they perceived as stressful, compared to 40% of university musicians. Despite pain catastrophizing correlating with depression, anxiety, low cognitive-functional abilities, depression was not a significant factor for pain catastrophizing in the model. In terms of music-specific variables, social identity as a musician was found to be a predictive variable however, it’s presence required a combination of various other variables to become a valid predictor.

## Discussion

To gain a better understanding of chronic pain experience in student musicians, this study investigated pain catastrophizing as one of the related pain behaviors and cognitions alongside seven complementary variables: anxiety, depression, depersonalisation, burnout, coping strategies, professional identity, and sleep. The results of the student musicians were compared to those reported from student non-musicians to identify any unique pathways to effect predicted by group membership.

Our tree model confirmed the bio-psycho-socio model from the chronic pain literature, which requires a combination of variables to cause a maladaptive process: anxiety, social identity, personal burnout, being over 29 years of age, low cognitive functional abilities, and burnout with fellow students. Group testing, however, showed that despite having perceived pain for significantly longer, music college musicians reported the lowest pain catastrophizing prevalence. In other words, pain catastrophizing was significantly lower in music college musicians compared to their university non-musician peers. Furthermore, music college musicians showed a significantly lower anxiety prevalence compared to university non-musicians. Finally, they reported higher job satisfaction and fewer missed workdays due to pain-based issues, when compared to their non-musician peers.

On the surface, these findings appear to disagree with current specialist literature on musicians’ health and wellbeing in that musicians’ pain significantly decreases their wellbeing (Leaver, Harris, and Palmer 2011). However, chronic pain, as previously stated, is not the prolongation of acute pain plus time, but a separate condition that is to be understood based on its bio-psycho-socio model. Briefly summarized this means that related pain behaviors and cognitions lead to substantial cortical plasticity alterations that could be viewed as ‘pain memories’, influencing painful and non-painful processing in the somato-sensory system. While pain in adolescence is generally under-researched, even more so in young musicians, Zamorano et al., (2023) have demonstrated distinct pain-related cortical plasticity changes in musicians. When considering the duration of the perceived pain in isolation, one could suppose that musicians would use maladaptive strategies that lead to such neurological changes. However, the long career of an average classical musician, combined with the high job satisfaction, especially in older musicians, call this hypothesis into question (Voltmer et al. 2012).

Before we consider more fully the role of professional identity as one possible variable, we need to re-consider basic pain management prognostics. There is reliable evidence that children with maladaptive coping strategies are more likely to perceive more pain when they reach adolescence. This trend increases even more from adolescence to adulthood (Kennedy et al 2008, Brattenberg 2004). In other words, as a group, it is highly unlikely that student musicians with maladaptive pain coping strategies might end up with a long career in music and high job satisfaction. While pain-induced beliefs and cognitions and neurological changes can be altered and even reversed, it is statistically unlikely to happen to an entire group. Furthermore, chronic pain processes are neither fast nor are they conscious decisions, but complex processes that grow and are reinforced over the course of a lifetime.

In this context, we take a closer look at musicians’ professional identity. In line with the relevant literature on chronic pain, our tree model underscored that social identity constitutes an important variable in the process of pain catastrophizing (Morley 2008). Music college musicians showed significantly higher scores in social and self-identity than university musicians. Additionally, they differed from the latter group by allocating more time to their daily practice. Both groups invested an equal number of years into their practical and theoretical musical training. Most instruments require years of practice to be mastered at (music) college level. We can therefore infer that both groups followed a similar education until reaching college or university level. Moreover, we can assume that during these formative training years young musicians got accustomed to a degree of performance-related pain. Professional identity not only reflects a person’s personal attitudes, but is also fed by the environment (Grove, Fish & Eklund 2004). Thus, their pain-enduring beliefs entered early during the process of musical training, and were, most probably, reinforced by parents, study friends, teachers and/or coaches.

At this point, we have no further biographical information on the details of this process, for instance the possible use of a no-pain-no-gain-method, or if pain-preventative measures were approached in training. We should thus endeavor, for now, not to attribute positive or negative implications to this pain management process. One conclusion we can draw is that perceived pain was not managed in a maladaptive way or that it impacted directly on their education related outcomes. Life goals, such as becoming a professional musician, were still pursued. Furthermore, we can draw the conclusion that the degree of higher education and/or place of study could make a difference for pain catastrophizing in student musicians. We know that both music college musicians and university musicians invested considerable time into the practice of their instruments/voice. For reasons of their own, they decided to enter music college, to follow a more academic path at a university, or to completely change their main study subject to something other than music. Their professional identity modified in line with these career choices, similarly to the professional identity of college athletes whose professional identity was found to decline after they chose a different career path to sports (Lally and Kerr 2013; Gembris, Heye, and Seifert 2018). It is probable, though speculative, that pain catastrophizing may have changed at this point. Further research is needed to understand the developmental aspects of this process more fully.

Two important variables that are frequently discussed in musician-specific literature on pain are depression and anxiety (Gembris, Heye, and Seifert 2018). The chronic pain literature describes both variables as having distinct properties that can lead to maladaptive pain perception processes. High anxiety is a reliable variable carrying considerable weight for maladaptive pain processes (Pfingsten et al. 2001). In our study, the university non-musicians showed a significantly higher anxiety prevalence compared to the two other groups, and consequently had the highest prevalence in pain catastrophizing.

A second variable, depression, did not carry such a significance. Music college musicians had significantly higher depression prevalence compared to university non-musicians yet showed significantly lower levels of pain catastrophizing. The reason for this might be that for depression to become a valid factor in pain catastrophizing it requires a specific point in time and degree of depression (e.g. mild). For instance, studies into chronic back pain found that depression only led to pain catastrophizing if the individuals had a mild depression (Kröner-Herwig et al. 2011). Finally, some studies on pain in musicians have presented the hypothesis that musicians experiences of pain can depend in part on their personality (Louven 2019). In contrast, chronic pain research points out that tying chronic pain to personality-based models should be most critically evaluated as there is little empirical evidence (Kröner-Herwig et al. 2011). Moreover, such an approach could lead to further stigmatization of individuals. In this context, personality-based models have a historical importance but going forward, more contemporary approaches using interpersonal models are more appropriate.

It is important to note the limitations of this study to allow a meaningful interpretation. Firstly, despite providing more than the number of participants required for meaningful results, the overall sample size is modest, which reflects the exploratory nature of the study. This could be addressed in future studies with larger numbers of participants. Secondly, and in keeping with the literature, some participants declined to disclose information about their mental health status (Clement et al. 2015). Thirdly, for anonymity reasons, variables such as gender were excluded. The inclusion of these variables might have provided some additional information, but on balance anonymity was deemed more important. Fourthly, this study was designed based on the empirical findings from studies in musicians’ pain perception. We did not anticipate that our findings would contrast with most of the current literature in musicians. In hindsight, it would have further elucidated our findings, had we included additional instruments for pain processing in the exploratory research design, to help interpret the data. We would suggest that future studies take this into account as they explore this area in more detail (e.g. development and degree of chronification or family history).

Overall, our findings provide several insights for chronic pain research in student musicians. Despite long-term perceived chronic pain (12 months and longer), music college musicians reported the best pain management strategies out of the three groups. Maladaptive strategies for chronic pain presented a larger problem for university non-musicians. This group showed the highest prevalence, increasing the likelihood of an overall negative long-term impact on their work, life, and wellbeing. The difference we found between music college musicians and university musicians in pain catastrophizing levels point to the conclusion that the environment shapes how chronic pain is managed. General pain chronification predictors all play a role, independent from whether the participant is a student musician or not. However, the depression variable requires a separate investigation based on its (time) specifics in chronic pain.

We conclude on the critical point that, whilst we found that music college musicians generally have good chronic pain management strategies, this should not minimize the fact that, as a group, they are disproportionately affected by chronic pain. Chronic pain management guidelines stipulate that, in contrast to the treatment of acute pain, a pain-free status may not be obtained (ASoA, 2010).

Moreover, pain control needs to be optimized and adverse outcomes minimized, such that quality of life may be enhanced. The implication for musicians is that prevention methods should be introduced at a young age as part of training. Chronic pain literature suggests that low scores in pain catastrophizing make individuals more open to pain preventative methods, as well as making compliance more likely (Kröner-Herwig et al. 2011). Hence a proactive and informed approach to early pain management during musical training, informed by the multiple predictive factors identified in this article, holds the promise for a future where at-risk musicians are identified at an earlier age and offered the strategies that will enhance their long-term career prospects and wellbeing.

## Data Availability

All data produced in the present study are available upon reasonable request to the authors

## Notes

### Competing Interest Statement

The authors have declared no competing interest.

### Funding Statement

This study did not receive any funding

### Author Declarations

Ethics committee of University of Sheffield (UK) gave ethical approval for this work

